# Mental health, HIV and healthcare-related discrimination among young sexual minority men in Kenya: implications for integrated care and stigma reduction

**DOI:** 10.64898/2026.09.12.26362875

**Authors:** Samuel W. Mwaniki, Richard M. Kilonzo, Peter M. Kaberia, Carlos K. Cheruiyot, Walter M. Nyagah, Humphrey K. Rop, Adrian D. Smith, Thesla Palanee-Phillips

**Affiliations:** Department of Health Services, Administration and Campus Support Services, University of Nairobi, Nairobi, Kenya; Alliance Care 360, Chicago, Illinois, USA; Data Science Program, African Population and Health Research Center, Dakar, Senegal; Department of Health Management and Informatics, School of Health Sciences, Kenyatta University, Nairobi, Kenya; Department of Epidemiology and Biostatistics, School of Public Health, Kwame Nkrumah University of Science and Technology, Kumasi, Ghana; Nuffield Department of Population Health, University of Oxford, Oxford, England; Wits Reproductive Health and HIV Institute, Faculty of Health Sciences, University of the Witwatersrand, Johannesburg, South Africa

## Abstract

Mental health conditions among young sexual minority men (YSMM) are a growing public health concern, yet they are understudied in sub-Saharan African settings. We aimed to estimate the prevalence of depressive and anxiety symptoms, and associated factors among YSMM in Kenya. Between February and March 2021, we conducted a cross-sectional bio-behavioral survey among YSMM aged ≥18 years enrolled in tertiary academic institutions in Nairobi, using respondent-driven sampling. Participants completed an electronically self-administered demographic and behavioral questionnaire on REDCap®. Depressive and anxiety symptoms were assessed using Patient Health Questionnaire-9 (PHQ-9) and Generalized Anxiety Disorder-7 (GAD-7) tools respectively, with scores ≥10 indicating moderate-to-severe symptoms. HIV antibody testing followed Kenyan national guidelines. Principal component analysis was used to derive stigma-related domains, and multivariate logistic regression identified factors associated with outcomes. Data analysis was done using Stata v.17 software. Among 242 participants, median age was 21 years (IQR 19 – 23). Prevalence of depressive, anxiety, co-occurring depressive/anxiety symptoms and HIV were: 23.7% (95% CI: 18.4 – 29.5), 16.9% (95% CI: 12.4 – 22.3), 12.4% (95% CI: 8.5 – 17.2) and 8.3% (95% CI: 5.1 – 12.5), respectively. Factors independently associated with depressive symptoms were: living with HIV (AOR: 4.88, 95% CI: 1.18 – 20.18, p = 0.029), moderate-to-severe anxiety symptoms (AOR: 25.63, 95% CI: 8.58 – 76.61, p <0.001), healthcare-related discrimination (AOR: 1.56, 95% CI: 1.07 – 2.28, p = 0.020) and early years of study (AOR: 2.79, 95% CI: 1.09 – 7.15, p = 0.033). Factors independently associated with anxiety symptoms were: moderate-to-severe depressive symptoms (AOR: 26.10, 95% CI: 8.10 – 84.15, p <0.001), attending a public academic institution (AOR: 3.33, 95% CI: 1.14 – 10.00, p = 0.028), and boarding school history (AOR: 4.35, 95% CI: 1.14 – 16.67, p = 0.032). YSMM in Nairobi experience a high burden of depressive and anxiety symptoms with substantial co-occurrence. Living with HIV, experiencing healthcare-related discrimination and educational context play key roles in shaping mental health outcomes among YSMM. These findings underscore the importance of integrating mental health with HIV services, alongside stigma-reduction interventions in both healthcare and educational settings, to improve access, quality of care, and outcomes for this young key population.

## Introduction

According to the Global Burden of Diseases, Injuries, and Risk Factors Study 2019, depressive and anxiety disorders were the two most prevalent mental disorders, ranking 13th and 24th respectively among the leading causes of health-related burden [1]. Although this 2019 study showed that the burden of depressive and anxiety disorders was high for both heterosexual females and males across many locations [1], there is evidence to suggest that sexual minority people may be at higher risk for mental disorders than their heterosexual counterparts [2]. The disproportionate burden of mental disorders among sexual minority people is present across all age groups. For instance, compared to heterosexual youth, sexual minority youth have three times higher odds of reporting depressive disorders [3].

Meyer’s minority stress model posits that stigma, prejudice, and discrimination create a stressful social environment that results in mental health problems for sexual minority individuals [4]. In keeping with other sexual minority populations, young sexual minority men aged 10 – 24 years (abbreviated hereafter as YSMM, and encompassing young gay, bisexual and other men who have sex with men) may experience internal stressors such as internalized homophobia, lack of self-acceptance, as well concealment of their sexual orientation in predominantly heteronormative and pervasively homophobic environments [5]. Within the vulnerability–stress model, mental disorders are understood to develop through the interaction between an individual’s innate susceptibility and external stressors [6]. For YSMM, external social stressors may include inadequate parental support [7], family-based victimization [8], school-based victimization [9] and gender role strain [10]. In countries where consensual adult same-sex practices are criminalized [11], the effect of these stressors may further be exacerbated by perpetration of violence against YSMM by various actors such as intimate partners, family members, religious leaders, police and strangers [12]. Additionally, available data suggest that YSMM experience higher rates of justice involvement through events like arrests and detainment, with each of these shown to be associated with greater odds of depression and anxiety symptoms [13]. Previously, YSMM have also reported experiences of stigma and discrimination in healthcare settings [14], factors which cause them to delay or completely avoid seeking care for mental health needs [15]. Overall, various factors within the minority stress and vulnerability–stress models mediate or account for the multifaceted relationship between sexual minority status and mental illnesses [16].

In the developmental phase of emerging adulthood (ages 18-25 years), individuals define their identities, cultivate values, forge new peer relationships, and explore newfound independence [17]. At the onset of this phase, many students typically transition from high school to college or university, entering a setting with less direct oversight of their behavior and more freedom [18]. Most students go through and complete their undergraduate studies while in this developmental stage. Navigating tasks such as self-exploration, evolving relationships and roles, and new expectations and experiences, often presents considerable challenges that may adversely affect the mental wellness of students [19]. Although the college transition and environment are generally stressful for most students, sexual minority students such as YSMM may face unique challenges due to an already existing higher risk of mental health disorders [20]. For instance, previous research findings in the US have shown that, compared to their heterosexual counterparts, sexual minority students consistently report higher levels of anxiety, negative feelings such as loneliness and sadness, suicide attempts, as well as poor academic performance [21]. Moreover, a study done in Nigeria showed that, in comparison to male heterosexual students, gay students were close to four times more likely to be depressed, and this was associated with internalized homophobia and expectations of stigma [22]. On the upside, college may present YSMM with the opportunity to freely meet and form friendships with peers of similar sexual orientations and behaviors. These new interactions may confer psychosocial benefits such as creating a sense of community, enabling the processes of self-acceptance, coming out and identity formation [23].

Although a number of studies have estimated the prevalence and correlates of mental health disorders among university students in sub-Saharan Africa, including Kenya [24–28], fewer studies have done this for YSMM students [22]. Furthermore, in the Kenyan context where homosexuality is criminalized and highly stigmatized, a number of studies have also investigated mental health disorders among sexual minority men [29–32], but to the best of our knowledge, none have specifically looked at YSMM in college/university settings. This study therefore aimed to fill this gap in knowledge, especially given the multiplicity of factors that predispose YSMM in college/university settings to mental health disorders. Accurate epidemiological data are essential to understand the burden and correlates of mental health disorders, and guide interventions that reduce risk and promote psychological wellbeing for YSMM attending college/university.

## Materials and Methods

The study methods are detailed in the published study protocol [33], and summarized below:

### Study design and setting

A cross-sectional bio-behavioral survey was conducted between February and March 2021, just after stringent COVID-19 prevention and control restrictions were eased in Kenya, and academic institutions allowed to resume in-person learning. Nairobi, Kenya’s capital, was selected due to its large population of tertiary students, with approximately 150 campuses of various universities and colleges located in the city and within its metropolis [34].

### Participants, sampling, and recruitment

YSMM were eligible to participate if they were willing and able to provide written informed consent for study participation, were aged ≥18 years, provided proof of registration as a student in a university or college in/within Nairobi, were assigned male sex at birth, and reported consensual receptive or insertive anal and/or oral sexual intercourse with another man in the last 12 months. As described in the detailed study protocol [33], the sample size was primarily calculated to estimate the prevalence of HIV and other sexually transmissible infections (STIs) among YSMM, based on WHO guidelines for bio-behavioral surveys among populations at higher risk of HIV infection [35]. Participants were enrolled using respondent-driven sampling (RDS) method, based on findings from formative qualitative research which showed that this sampling method was appropriate and acceptable for recruiting YSMM into research [36]. In summary, RDS is a chain referral sampling method that integrates a mathematical model that weights the sample to compensate for the non-random sampling and minimizes traditional snowballing bias [23]. To begin the recruitment, 6 seeds were selected from the YSMM who took part in the formative research. Each seed and subsequent participants were issued with 3 coupons to help recruit their peers, until the survey had recruited 120 participants (inclusive of seeds), then 2 coupons until the 150th participant, after which no more coupons were issued. Each participant was reimbursed 1000 Kenyan shillings (equivalent to US$ 10 at the time) for their time and expenses related to travel to and from the study site, and 300 Kenyan shillings (equivalent to US$ 3 at the time) for every peer they recruited into the study. A more detailed account of the recruitment process, as well as the results of the primary objectives of the study are reported elsewhere [37–38].

### Data collection tools and procedures

#### Outcome variables

The outcome variables, namely: depression and anxiety symptoms were measured using respective psychometric scales. Depressive symptoms were assessed using the Patient Health Questionnaire-9 (PHQ-9) scale [39], applying a cut-off score of ≥10 for moderate-to-severe symptoms [32]. As a screening tool for depression, the PHQ-9 scale has performed well in terms of internal validity and reliability in previous studies with Kenyan SMM [29–32], as well as university students in Nigeria [40]. Anxiety symptoms were assessed using the Generalized Anxiety Disorders-7 (GAD-7) [41], with a cut-off score of ≥10 for moderate-to-severe symptoms [42]. The GAD-7 scale has been validated among adults living with HIV in Kenya [43], and shown good internal validity when used with university students in Ethiopia [44]. Both the PHQ-9 and GAD-7 scales showed good internal validity for our study sample (Cronbach’s alpha coefficient = 0.73).

#### Exposure variables

As per the study protocol [33], data on exposure variables were collected under the following domains: sociodemographic characteristics, sexual behavior history, contextual factors such as disclosure of same-sex attraction to family and service providers, and test results for HIV and any of five curable STI agents (*Chlamydia trachomatis, Mycoplasma genitalium, Neisseria gonorrhea, Treponema pallidum* and/or *Trichomonas vaginalis)* at the time of the study. Perceived stress was assessed using the Perceived Stress Scale-10 (PSS-10) [45], with a cut-off score of ≥14 for moderate-to-high stress. The PSS-10 has been validated among Ethiopian university students [46], and used to assess perceived stress among YSMM college students in the US [47]. Alcohol use was measured using the Alcohol Use Disorders Identification Test (AUDIT) [48], which has previously been used to assess alcohol use among university students in Kenya [49], as well as YSMM in the US [50]. Participants were also asked about use of other injectable and non-injectable drugs for non-medical reasons. Experiences of stigma, discrimination and violence in various settings were assessed using cross-culturally relevant metrics for SMM across Sub-Saharan countries, as characterized by Augustinavicius *et al*. [51]. The behavioral survey was conducted through self-administered interviews on tablets using a questionnaire on REDCap® software (Vanderbilt University, TN, USA). The questionnaire was administered in English which is the language of instruction in Kenyan academic tertiary institutions.

### Data analysis

Data analysis excluded six seeds who were purposely selected to begin the RDS recruitment. The outcome variables of depression and anxiety were both dichotomized at scores of 0-9 (none to mild) and ≥10 (moderate-to-severe). Descriptive statistics were computed using measures of central tendency and variability, frequencies and proportions. Some continuous variables such as age and self-reported number of sex partners were converted into binary categories and analyzed as such. The chi-square (χ²) test was used to determine whether there were significant differences between participants with and without the outcome variables, by probable exposure variables.

Exploratory principal component analysis (PCA) was conducted on 19 stigma-, discrimination- and violence-related items [51], to reduce redundancy and identify underlying dimensions of these experiences. Sampling adequacy for PCA was assessed using the Kaiser–Meyer–Olkin (KMO) measure, and factorability was confirmed using Bartlett’s test of sphericity. Components were extracted using principal-component factoring, and the number of components retained was guided by the Kaiser criterion (Eigenvalues > 1). To enhance interpretability, orthogonal varimax rotation was applied. Items with rotated factor loadings ≥0.50 were considered to load meaningfully onto a component. Component labels were assigned inductively based on thematic similarity among items loading on each component, rather than pre-specified theoretical constructs. Component scores were generated and used as exposure variables in subsequent multivariate logistic regression analyses.

Multivariate logistic regression models were used to measure associations between various exposure variables (including components derived from PCA of stigma, discrimination and violence items), and prevalence of depressive and anxiety symptoms. All exposure variables with a p < 0.2 in bivariate analysis and PCA components with Eigenvalues > 1 were included in the multivariate model. Both crude odds ratios (COR) and adjusted odds ratios (AOR) alongside their corresponding 95% confidence intervals (CI) and p-values were calculated for each exposure variable and PCA component in the final model. Variables and PCA components with p-value <0.05 were considered statistically significant. All statistical tests were two-sided. To assess multicollinearity among the exposure variables in the final model, Variance Inflation Factors (VIFs) were computed. Model calibration was assessed using the Hosmer–Lemeshow goodness-of-fit test. Data analysis was done using Stata version 17 (College Station, TX: StataCorp LLC).

### Ethical considerations

This study was approved by University of the Witwatersrand Human Research Ethics Committee-Medical (Ref. No. M200215) and University of Nairobi-Kenyatta National Hospital Ethics and Research Committee (Ref. No. P990/12/2019). All study procedures were carried out in line with the principles of the Declaration of Helsinki. All participants provided written informed consent.

## Results

### Participant characteristics

The characteristics of the participants in total sample (242) and by depressive and anxiety symptoms are shown in Tables 1 and 2, respectively. Median age was 21 years (interquartile range[IQR], 19–23). A majority of the participants (90.5%) identified as cisgender male, close to two-thirds (63.2%) as gay, one-third (33.2%) as bisexual, and a small proportion (3.7%) as heterosexual. More than half (58.3%) attended university, close to three-quarters (71.9%) were in public academic institutions, and four-fifths (79.3%) resided away from their families. Almost two-thirds (64.5%) were taking science, technology, engineering and mathematics-related courses, with almost a similar proportion (65.8%) in early years of college/university. In terms of sexual behaviors, one-third (33.0%) had their sexual debut with a man before the age of 18 years. Almost three-quarters (71.4%) had more than one sex partner in the last 12 months, with a small proportion (16.0%) having participated in group sex within the same period. Contextually, only a few participants had disclosed same-sex attraction to family members (5.4%), healthcare providers (14.9%), or college/university counselors (8.7%). Prevalence of HIV and at least one of five curable STIs were 8.3% and 64.5%, respectively. Varying proportions of participants had anticipated and/or experienced stigma, discrimination, and/or violence across the 19 items assessed, with feeling afraid to seek health services (29.3%) and being denied restaurant service (3.7%) the highest and lowest proportions, respectively.

**Table 1:**
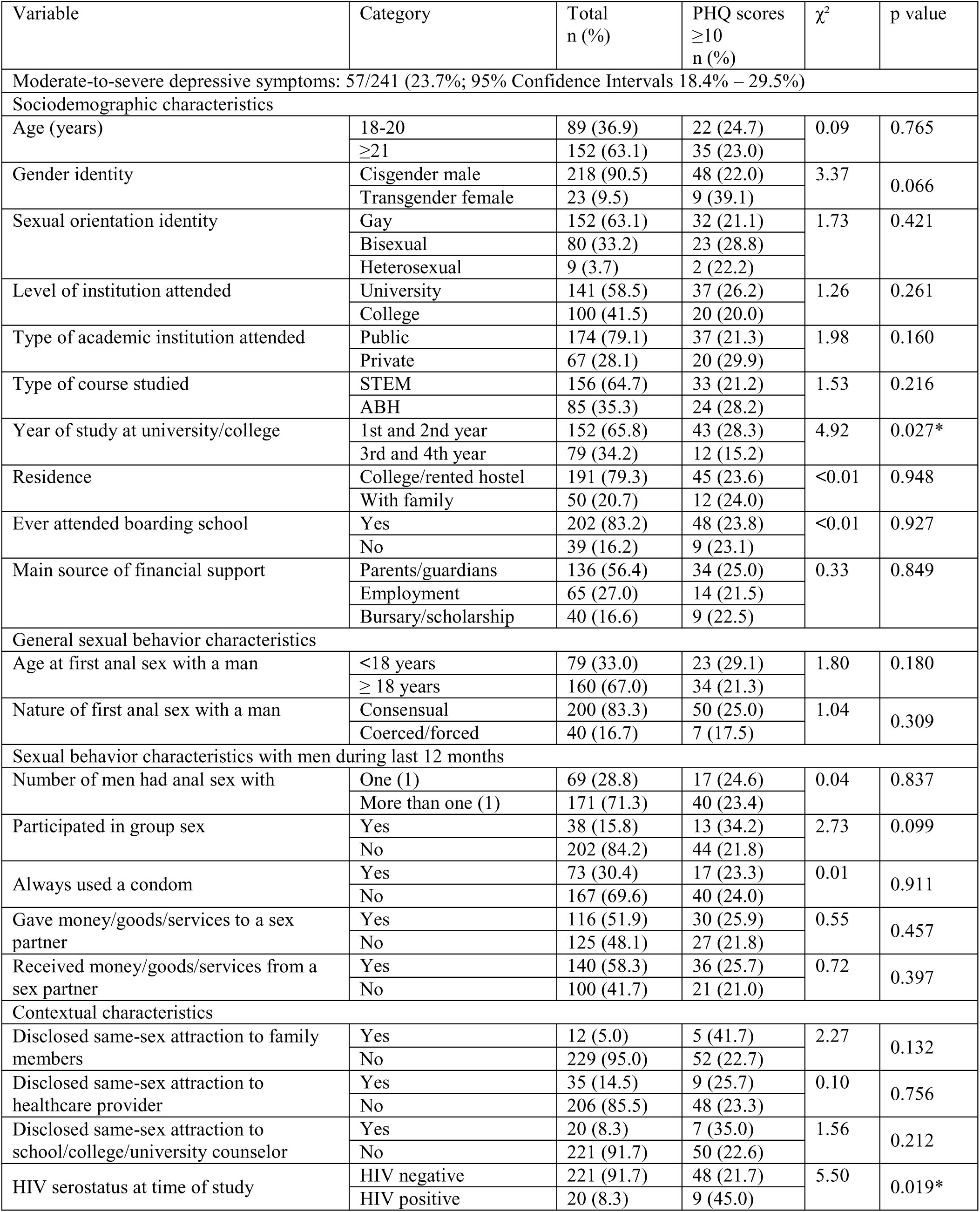
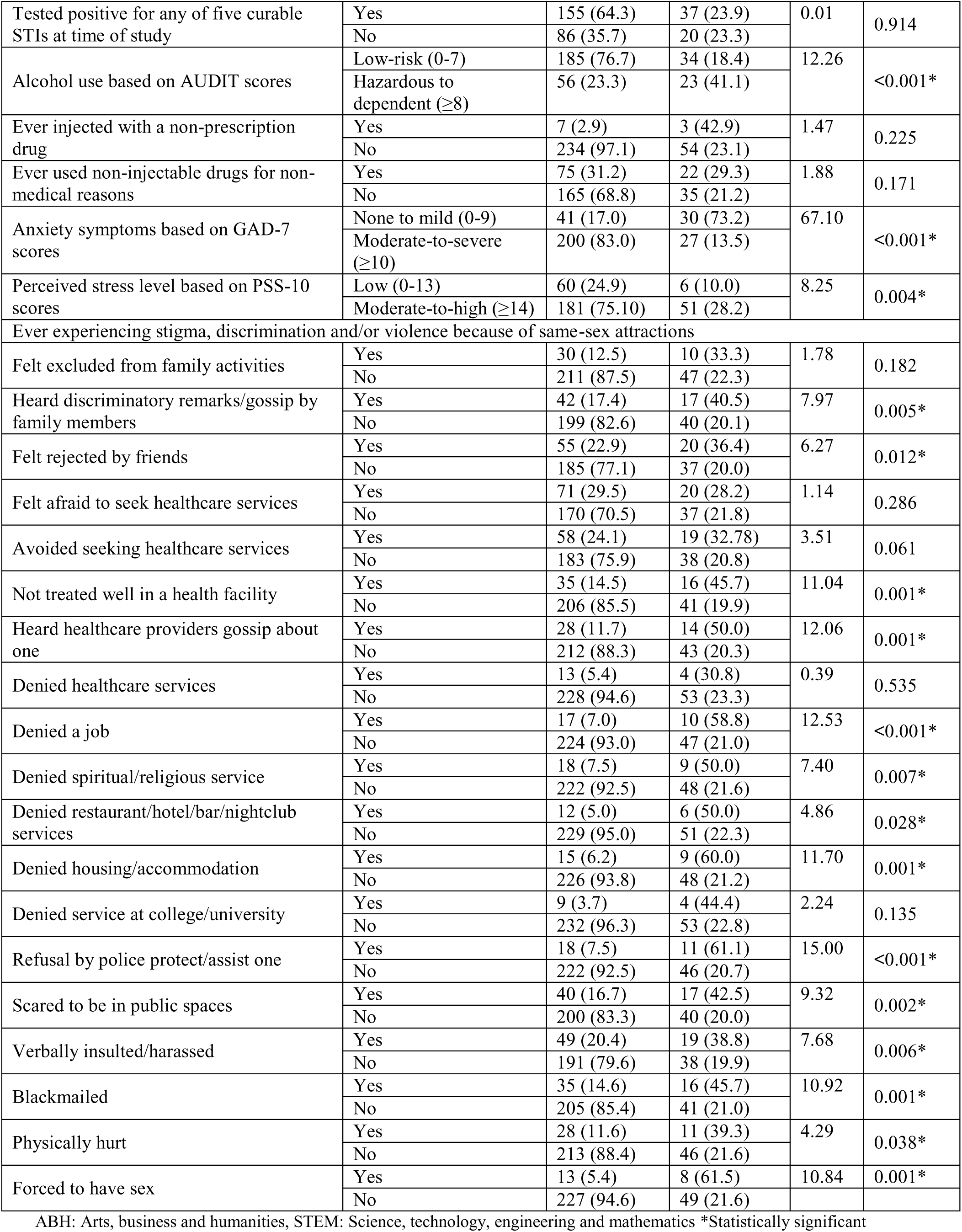
Participant characteristics by total and level of depressive symptoms (n=241)

| Variable | Category | Total<br>n (%) | PHQ scores<br>≥10<br>n (%) | χ<sup>2</sup> | p value |
| --- | --- | --- | --- | --- | --- |
| Moderate-to-severe depressive symptoms: 57/241 (23.7%; 95% Confidence Intervals 18.4% – 29.5%) |  |  |  |  |  |
| Sociodemographic characteristics |  |  |  |  |  |
| Age (years) | 18-20 | 89 (36.9) | 22 (24.7) | 0.09 | 0.765 |
|  | ≥21 | 152 (63.1) | 35 (23.0) |  |  |
| Gender identity | Cisgender male | 218 (90.5) | 48 (22.0) | 3.37 | 0.066 |
|  | Transgender female | 23 (9.5) | 9 (39.1) |  |  |
| Sexual orientation identity | Gay | 152 (63.1) | 32 (21.1) | 1.73 | 0.421 |
|  | Bisexual | 80 (33.2) | 23 (28.8) |  |  |
|  | Heterosexual | 9 (3.7) | 2 (22.2) |  |  |
| Level of institution attended | University | 141 (58.5) | 37 (26.2) | 1.26 | 0.261 |
|  | College | 100 (41.5) | 20 (20.0) |  |  |
| Type of academic institution attended | Public | 174 (79.1) | 37 (21.3) | 1.98 | 0.160 |
|  | Private | 67 (28.1) | 20 (29.9) |  |  |
| Type of course studied | STEM | 156 (64.7) | 33 (21.2) | 1.53 | 0.216 |
|  | ABH | 85 (35.3) | 24 (28.2) |  |  |
| Year of study at university/college | 1st and 2nd year | 152 (65.8) | 43 (28.3) | 4.92 | 0.027* |
|  | 3rd and 4th year | 79 (34.2) | 12 (15.2) |  |  |
| Residence | College/rented hostel | 191 (79.3) | 45 (23.6) | <0.01 | 0.948 |
|  | With family | 50 (20.7) | 12 (24.0) |  |  |
| Ever attended boarding school | Yes | 202 (83.2) | 48 (23.8) | <0.01 | 0.927 |
|  | No | 39 (16.2) | 9 (23.1) |  |  |
| Main source of financial support | Parents/guardians | 136 (56.4) | 34 (25.0) | 0.33 | 0.849 |
|  | Employment | 65 (27.0) | 14 (21.5) |  |  |
|  | Bursary/scholarship | 40 (16.6) | 9 (22.5) |  |  |
| General sexual behavior characteristics |  |  |  |  |  |
| Age at first anal sex with a man | <18 years | 79 (33.0) | 23 (29.1) | 1.80 | 0.180 |
|  | ≥ 18 years | 160 (67.0) | 34 (21.3) |  |  |
| Nature of first anal sex with a man | Consensual | 200 (83.3) | 50 (25.0) | 1.04 | 0.309 |
|  | Coerced/forced | 40 (16.7) | 7 (17.5) |  |  |
| Sexual behavior characteristics with men during last 12 months |  |  |  |  |  |
| Number of men had anal sex with | One (1) | 69 (28.8) | 17 (24.6) | 0.04 | 0.837 |
|  | More than one (1) | 171 (71.3) | 40 (23.4) |  |  |
| Participated in group sex | Yes | 38 (15.8) | 13 (34.2) | 2.73 | 0.099 |
|  | No | 202 (84.2) | 44 (21.8) |  |  |
| Always used a condom | Yes | 73 (30.4) | 17 (23.3) | 0.01 | 0.911 |
|  | No | 167 (69.6) | 40 (24.0) |  |  |
| Gave money/goods/services to a sex partner | Yes | 116 (51.9) | 30 (25.9) | 0.55 | 0.457 |
|  | No | 125 (48.1) | 27 (21.8) |  |  |
| Received money/goods/services from a sex partner | Yes | 140 (58.3) | 36 (25.7) | 0.72 | 0.397 |
|  | No | 100 (41.7) | 21 (21.0) |  |  |
| Contextual characteristics |  |  |  |  |  |
| Disclosed same-sex attraction to family members | Yes | 12 (5.0) | 5 (41.7) | 2.27 | 0.132 |
|  | No | 229 (95.0) | 52 (22.7) |  |  |
| Disclosed same-sex attraction to healthcare provider | Yes | 35 (14.5) | 9 (25.7) | 0.10 | 0.756 |
|  | No | 206 (85.5) | 48 (23.3) |  |  |
| Disclosed same-sex attraction to school/college/university counselor | Yes | 20 (8.3) | 7 (35.0) | 1.56 | 0.212 |
|  | No | 221 (91.7) | 50 (22.6) |  |  |
| HIV serostatus at time of study | HIV negative | 221 (91.7) | 48 (21.7) | 5.50 | 0.019* |
|  | HIV positive | 20 (8.3) | 9 (45.0) |  |  |
| Tested positive for any of five curable STIs at time of study | Yes | 155 (64.3) | 37 (23.9) | 0.01 | 0.914 |
|  | No | 86 (35.7) | 20 (23.3) |  |  |
| Alcohol use based on AUDIT scores | Low-risk (0-7) | 185 (76.7) | 34 (18.4) | 12.26 | <0.001* |
|  | Hazardous to dependent (≥8) | 56 (23.3) | 23 (41.1) |  |  |
| Ever injected with a non-prescription drug | Yes | 7 (2.9) | 3 (42.9) | 1.47 | 0.225 |
|  | No | 234 (97.1) | 54 (23.1) |  |  |
| Ever used non-injectable drugs for non-medical reasons | Yes | 75 (31.2) | 22 (29.3) | 1.88 | 0.171 |
|  | No | 165 (68.8) | 35 (21.2) |  |  |
| Anxiety symptoms based on GAD-7 scores | None to mild (0-9) | 41 (17.0) | 30 (73.2) | 67.10 | <0.001* |
|  | Moderate-to-severe (≥10) | 200 (83.0) | 27 (13.5) |  |  |
| Perceived stress level based on PSS-10 scores | Low (0-13) | 60 (24.9) | 6 (10.0) | 8.25 | 0.004* |
|  | Moderate-to-high (≥14) | 181 (75.10) | 51 (28.2) |  |  |
| Ever experiencing stigma, discrimination and/or violence because of same-sex attractions |  |  |  |  |  |
| Felt excluded from family activities | Yes | 30 (12.5) | 10 (33.3) | 1.78 | 0.182 |
|  | No | 211 (87.5) | 47 (22.3) |  |  |
| Heard discriminatory remarks/gossip by family members | Yes | 42 (17.4) | 17 (40.5) | 7.97 | 0.005* |
|  | No | 199 (82.6) | 40 (20.1) |  |  |
| Felt rejected by friends | Yes | 55 (22.9) | 20 (36.4) | 6.27 | 0.012* |
|  | No | 185 (77.1) | 37 (20.0) |  |  |
| Felt afraid to seek healthcare services | Yes | 71 (29.5) | 20 (28.2) | 1.14 | 0.286 |
|  | No | 170 (70.5) | 37 (21.8) |  |  |
| Avoided seeking healthcare services | Yes | 58 (24.1) | 19 (32.78) | 3.51 | 0.061 |
|  | No | 183 (75.9) | 38 (20.8) |  |  |
| Not treated well in a health facility | Yes | 35 (14.5) | 16 (45.7) | 11.04 | 0.001* |
|  | No | 206 (85.5) | 41 (19.9) |  |  |
| Heard healthcare providers gossip about one | Yes | 28 (11.7) | 14 (50.0) | 12.06 | 0.001* |
|  | No | 212 (88.3) | 43 (20.3) |  |  |
| Denied healthcare services | Yes | 13 (5.4) | 4 (30.8) | 0.39 | 0.535 |
|  | No | 228 (94.6) | 53 (23.3) |  |  |
| Denied a job | Yes | 17 (7.0) | 10 (58.8) | 12.53 | <0.001* |
|  | No | 224 (93.0) | 47 (21.0) |  |  |
| Denied spiritual/religious service | Yes | 18 (7.5) | 9 (50.0) | 7.40 | 0.007* |
|  | No | 222 (92.5) | 48 (21.6) |  |  |
| Denied restaurant/hotel/bar/nightclub services | Yes | 12 (5.0) | 6 (50.0) | 4.86 | 0.028* |
|  | No | 229 (95.0) | 51 (22.3) |  |  |
| Denied housing/accommodation | Yes | 15 (6.2) | 9 (60.0) | 11.70 | 0.001* |
|  | No | 226 (93.8) | 48 (21.2) |  |  |
| Denied service at college/university | Yes | 9 (3.7) | 4 (44.4) | 2.24 | 0.135 |
|  | No | 232 (96.3) | 53 (22.8) |  |  |
| Refusal by police protect/assist one | Yes | 18 (7.5) | 11 (61.1) | 15.00 | <0.001* |
|  | No | 222 (92.5) | 46 (20.7) |  |  |
| Scared to be in public spaces | Yes | 40 (16.7) | 17 (42.5) | 9.32 | 0.002* |
|  | No | 200 (83.3) | 40 (20.0) |  |  |
| Verbally insulted/harassed | Yes | 49 (20.4) | 19 (38.8) | 7.68 | 0.006* |
|  | No | 191 (79.6) | 38 (19.9) |  |  |
| Blackmailed | Yes | 35 (14.6) | 16 (45.7) | 10.92 | 0.001* |
|  | No | 205 (85.4) | 41 (21.0) |  |  |
| Physically hurt | Yes | 28 (11.6) | 11 (39.3) | 4.29 | 0.038* |
|  | No | 213 (88.4) | 46 (21.6) |  |  |
| Forced to have sex | Yes | 13 (5.4) | 8 (61.5) | 10.84 | 0.001* |
|  | No | 227 (94.6) | 49 (21.6) |  |  |

ABH: Arts, business and humanities, STEM: Science, technology, engineering and mathematics \*Statistically significant

**Table 2:** Participant characteristics by total and level of anxiety symptoms (n=242)

| Variable | Category | Total<br>n (%) | GAD<br>scores ≥10<br>n (%) | χ<sup>2</sup> | p value |
| --- | --- | --- | --- | --- | --- |
| Moderate-to-severe anxiety symptoms: 41/242 (16.9%; 95% Confidence Intervals 12.4% – 22.3%) |  |  |  |  |  |
| Sociodemographic characteristics |  |  |  |  |  |
| Age (years) | 18-20 | 89 (36.8) | 21 (23.6) | 4.43 | 0.035* |
|  | ≥21 | 153 (63.2) | 20 (13.1) |  |  |
| Gender identity | Cisgender male | 219 (90.5) | 35 (16.0) | 1.51 | 0.219 |
|  | Transgender female | 23 (9.5) | 6 (26.1) |  |  |
| Sexual orientation identity | Gay | 153 (63.2) | 23 (15.0) | 1.11 | 0.575 |
|  | Bisexual | 80 (33.1) | 16 (20.0) |  |  |
|  | Heterosexual | 9 (3.7) | 2 (22.2) |  |  |
| Level of institution attended | University | 141 (58.3) | 29 (20.6) | 3.15 | 0.076 |
|  | College | 101 (41.7) | 11 (11.9) |  |  |
| Type of academic institution attended | Public | 174 (71.9) | 21 (12.1) | 10.45 | 0.001* |
|  | Private | 68 (28.1) | 20 (29.4) |  |  |
| Type of course studied | STEM | 156 (64.5) | 24 (15.4) | 0.76 | 0.384 |
|  | ABH | 86 (35.5) | 17(19.8) |  |  |
| Year of study at university/college | 1st and 2nd year | 153 (66.0) | 31 (20.3) | 2.07 | 0.150 |
|  | 3rd and 4th year | 79 (34.0) | 10 (12.7) |  |  |
| Residence | College/rented hostel | 192 (79.3) | 28 (14.6) | 3.67 | 0.055 |
|  | With family | 50 (20.7) | 13 (26.0) |  |  |
| Ever attended boarding school | Yes | 203 (83.9) | 31 (15.3) | 2.50 | 0.114 |
|  | No | 39 (16.1) | 10 (25.6) |  |  |
| Main source of financial support | Parents/guardians | 137 (56.6) | 26 (19.0) | 0.95 | 0.621 |
|  | Employment | 65 (26.9) | 9 (13.9) |  |  |
|  | Bursary/scholarship | 40 (16.5) | 6 (15.0) |  |  |
| General sexual behavior characteristics |  |  |  |  |  |
| Age at first anal sex with a man | <18 years | 79 (33.0) | 17 (21.5) | 1.64 | 0.201 |
|  | ≥ 18 years | 161 (67.0) | 24 (14.9) |  |  |
| Nature of first anal sex with a man | Consensual | 201 (83.4) | 36 (17.9) | 0.69 | 0.406 |
|  | Coerced/forced | 40 (16.6) | 5 (12.5) |  |  |
| Sexual behavior characteristics with men during last 12 months |  |  |  |  |  |
| Number of men had anal sex with | One (1) | 69 (28.6) | 11 (16.0) | 0.08 | 0.779 |
|  | More than one (1) | 172 (71.4) | 30 (17.4) |  |  |
| Participated in group sex | Yes | 39 (16.2) | 5 (12.8) | 0.58 | 0.447 |
|  | No | 202 (83.8) | 36 (17.8) |  |  |
| Always used a condom | Yes | 74 (30.7) | 11 (14.9) | 0.35 | 0.555 |
|  | No | 167 (69.3) | 30 (18.0) |  |  |
| Gave money/goods/services to a sex partner | Yes | 116 (51.9) | 16 (13.8) | 1.65 | 0.200 |
|  | No | 125 (48.1) | 25 (20.0) |  |  |
| Received money/goods/services from a sex partner | Yes | 141 (58.5) | 21 (14.9) | 1.08 | 0.299 |
|  | No | 100 (41.5) | 20 (20.0) |  |  |
| Contextual characteristics |  |  |  |  |  |
| Disclosed same-sex attraction to family members | Yes | 13 (5.4) | 4 (30.8) | 1.87 | 0.172 |
|  | No | 229 (94.6) | 37 (16.2) |  |  |
| Disclosed same-sex attraction to healthcare provider | Yes | 36 (14.9) | 10 (27.8) | 3.53 | 0.060 |
|  | No | 206 (85.1) | 31 (15.1) |  |  |
| Disclosed same-sex attraction to school/college/university counselor | Yes | 21 (8.7) | 5 (23.8) | 0.77 | 0.380 |
|  | No | 221 (91.3) | 36 (16.3) |  |  |
| HIV serostatus at time of study | HIV negative | 222 (91.7) | 35 (15.8) | 2.64 | 0.104 |
|  | HIV positive | 20 (8.3) | 6 (30.0) |  |  |
| Tested positive for any of five curable STIs at time of study | Yes | 156 (64.5) | 26 (16.7) | 0.02 | 0.878 |
|  | No | 86 (35.5) | 15 (17.4) |  |  |
| Alcohol use based on AUDIT scores | Low-risk (0-7) | 185 (76.5) | 28 (15.1) | 1.82 | 0.177 |
|  | Hazardous to dependent (≥8) | 57 (23.5) | 13 (22.8) |  |  |
| Ever injected with a non-prescription drug | Yes | 7 (2.9) | 1 (14.3) | 0.04 | 0.849 |
|  | No | 235 (97.1) | 40 (17.0) |  |  |
| Ever used non-injectable drugs for non-medical reasons | Yes | 75 (31.1) | 13 (17.3) | 0.04 | 0.837 |
|  | No | 166 (68.9) | 27 (16.3) |  |  |
| Depressive symptoms based on PHQ-9 scores | None to mild (0-9) | 184 (76.4) | 11 (6.0) | 67.10 | <0.001* |
|  | Moderate-to-severe (≥10) | 57 (23.6) | 30 (52.6) |  |  |
| Perceived stress level based on PSS-10 scores | Low (0-13) | 61 (25.2) | 2 (3.3) | 10.82 | 0.001* |
|  | Moderate-to-high (≥14) | 181 (74.8) | 39 (21.6) |  |  |
| Ever experiencing stigma, discrimination and/or violence because of same-sex attractions |  |  |  |  |  |
| Felt excluded from family activities | Yes | 30 (12.4) | 7 (23.3) | 0.99 | 0.319 |
|  | No | 212 (87.6) | 34 (16.0) |  |  |
| Heard discriminatory remarks/gossip by family members | Yes | 42 (17.4) | 10 (23.8) | 1.70 | 0.192 |
|  | No | 200 (82.6) | 31 (15.5) |  |  |
| Felt rejected by friends | Yes | 55 (22.8) | 16 (29.1) | 7.36 | 0.007* |
|  | No | 186 (77.2) | 25 (13.4) |  |  |
| Felt afraid to seek healthcare services | Yes | 71 (29.3) | 17 (23.9) | 3.50 | 0.061 |
|  | No | 171 (70.7) | 24 (14.0) |  |  |
| Avoided seeking healthcare services | Yes | 58 (24.0) | 16 (27.6) | 6.14 | 0.013* |
|  | No | 184 (76.0) | 25 (13.6) |  |  |
| Not treated well in a health facility | Yes | 36 (14.9) | 11 (30.6) | 5.57 | 0.018* |
|  | No | 206 (85.1) | 30 (14.6) |  |  |
| Heard healthcare providers gossip about one | Yes | 28 (11.6) | 6 (21.4) | 0.44 | 0.508 |
|  | No | 213 (88.4) | 35 (16.4) |  |  |
| Denied healthcare services | Yes | 13 (5.4) | 3 (23.1) | 0.37 | 0.644 |
|  | No | 229 (94.6) | 38 (16.6) |  |  |
| Denied a job | Yes | 17 (7.0) | 6 (35.3) | 4.38 | 0.036* |
|  | No | 225 (93.0) | 35 (15.6) |  |  |
| Denied spiritual/religious service | Yes | 18 (7.5) | 7 (38.9) | 6.60 | 0.010* |
|  | No | 223 (92.5) | 34 (15.3) |  |  |
| Denied restaurant/hotel/bar/nightclub services | Yes | 12 (5.0) | 4 (33.3) | 2.41 | 0.121 |
|  | No | 230 (95.0) | 37 (16.1) |  |  |
| Denied housing/accommodation | Yes | 15 (6.2) | 7 (46.7) | 10.04 | 0.002* |
|  | No | 227 (93.8) | 34 (15.0) |  |  |
| Denied service at college/university | Yes | 9 (3.7) | 1 (11.1) | 0.23 | 0.635 |
|  | No | 233 (96.3) | 40 (17.2) |  |  |
| Refusal by police protect/assist one | Yes | 18 (7.5) | 8 (44.4) | 10.37 | 0.001* |
|  | No | 223 (92.5) | 33 (14.8) |  |  |
| Scared to be in public spaces | Yes | 40 (16.6) | 16 (40.0) | 17.95 | <0.001* |
|  | No | 201 (83.4) | 25 (12.4) |  |  |
| Verbally insulted/harassed | Yes | 50 (20.7) | 18 (36.0) | 16.11 | <0.001* |
|  | No | 191 (79.3) | 23 (12.0) |  |  |
| Blackmailed | Yes | 35 (14.5) | 11 (31.4) | 6.03 | 0.014* |
|  | No | 206 (85.5) | 30 (14.6) |  |  |
| Physically hurt | Yes | 28 (11.6) | 7 (25.0) | 1.46 | 0.227 |
|  | No | 214 (88.4) | 34 (15.9) |  |  |
| Forced to have sex | Yes | 13 (5.4) | 6 (46.2) | 8.67 | 0.003* |
|  | No | 228 (94.6) | 34 (14.9) |  |  |

ABH: Arts, business and humanities, STEM: Science, technology, engineering and mathematics \*Statistically significant

### Prevalence and correlates of depressive and anxiety symptoms

Based on a PHQ-9 cut-off score of ≥10, the prevalence of moderate-to-severe depressive symptoms was 57/241 (23.7%; 95% CI: 18.4%–29.5%). Based on a GAD-7 cut-off score of ≥10, the prevalence of moderate-to-severe anxiety symptoms was 41/242 (16.9%; 95% CI: 12.4%– 22.3%). Co-occurring moderate-to-severe depressive and anxiety symptoms were present in 30/241 participants (12.4%; 95% CI: 8.5 – 17.2).

As shown in Table 1, in bivariate analyses, the prevalence of moderate-to-severe depressive symptoms was significantly higher among participants in their first and second year than those in their third and fourth year of study (χ² = 4.92, p = 0.027), participants living with HIV than those without (χ² = 5.50, p = 0.019), those with hazardous to dependent alcohol use than those with low-risk alcohol use (χ² = 12.26, p < 0.001), participants exhibiting moderate-to-severe anxiety symptoms than those with none to mild anxiety symptoms (χ² = 67.10, p < 0.001), and those with moderate-to-high perceived stress than those with low perceived stress (χ² = 8.25, p = 0.004). Participants reporting exposure to 14 of the 19 stigma-, discrimination-, and violence-related items also had a significantly higher prevalence of moderate-to-severe depressive symptoms compared with those reporting no exposure to the respective items (all p < 0.05).

The results of the bivariate analyses for anxiety symptoms are shown in Table 2. The prevalence of moderate-to-severe anxiety symptoms was significantly higher among participants aged 18–20 years compared with those aged 21 years and above (χ² = 4.43, p = 0.035), participants in private academic institutions compared with those in public academic institutions (χ² = 10.45, p = 0.001), participants exhibiting moderate-to-severe depressive symptoms compared with those with none to mild depressive symptoms (χ² = 67.10, p < 0.001), and those with moderate-to-high perceived stress compared with those with low perceived stress (χ² = 10.82, p = 0.001).

Participants reporting exposure to 11 of the 19 stigma-, discrimination-, and violence-related items also had a significantly higher prevalence of moderate-to-severe anxiety symptoms compared with those reporting no exposure to the respective items (all p < 0.05).

The findings of the exploratory principal component analysis (PCA) of the 19 stigma-, discrimination-, and violence-related items are shown in Table 3. The PCA yielded five components with Eigenvalues greater than one, together accounting for 59.4% of the total variance. The overall Kaiser–Meyer–Olkin (KMO) measure of sampling adequacy was 0.83, indicating excellent suitability for factor analysis, and Bartlett’s test of sphericity was significant (χ² (171) = 1491.46, p < 0.001). Examination of the rotated solution revealed a clear and interpretable structure. The first component comprised items reflecting community or institutional-level exclusionary practices. The second component included items capturing differential or unfair treatment in healthcare settings. The third component reflected negative social response in family and religious settings, whereas the fourth component represented anticipated negative social response in healthcare and social settings. The fifth component consisted of items reflecting fear and experience of violence. Most items loaded strongly and uniquely onto a single component. The five components with Eigenvalues greater than one were subsequently included as predictors in multivariate logistic regression analyses.

**Table 3:** Exploratory principal component analysis (PCA) with varimax rotation of stigma, discrimination and violence items (loading ≥ 0.50)

| Stigma, discrimination, and violence items | Factors |  |  |  |  |
| --- | --- | --- | --- | --- | --- |
|  | 1 | 2 | 3 | 4 | 5 |
| Denied housing/accommodation | 0.82 |  |  |  |  |
| Denied restaurant/hotel/bar/nightclub services | 0.73 |  |  |  |  |
| Refusal by police protect/assist one | 0.72 |  |  |  |  |
| Denied service at college/university | 0.52 |  |  |  |  |
| Not treated well in a health facility |  | 0.72 |  |  |  |
| Heard healthcare providers gossip about one |  | 0.72 |  |  |  |
| Denied healthcare services |  | 0.60 |  |  |  |
| Felt excluded from family activities |  |  | 0.69 |  |  |
| Heard discriminatory remarks by family members |  |  | 0.64 |  |  |
| Denied spiritual/religious service |  |  | 0.58 |  |  |
| Felt afraid to seek healthcare services |  |  |  | 0.85 |  |
| Avoided seeking healthcare services |  |  |  | 0.79 |  |
| Felt rejected by friends |  |  |  | 0.54 |  |
| Scared to be in public spaces |  |  |  |  | 0.76 |
| Verbally insulted/harassed |  |  |  |  | 0.72 |

Inductively assigned labels of the factors based on item content: Factor 1: Experienced community/institutional-level discrimination, Factor 2: Experienced healthcare-related discrimination, Factor 3: Perceived stigma and experienced discrimination in family/religious settings, Factor 4: Anticipated stigma in healthcare/social settings, Factor 5: Fear and experience of violence

The findings of the multivariate logistic regression analyses are shown in Tables 4 and 5. As shown in Table 4, independent factors significantly associated with moderate-to-severe depressive symptoms were being in the first or second year of study (AOR: 2.79, 95% CI: 1.09– 7.15, p = 0.033), living with HIV (AOR: 4.88, 95% CI: 1.18–20.18, p = 0.029), having moderate-to-severe anxiety symptoms (AOR: 25.63, 95% CI: 8.58–76.61, p < 0.001), and experiencing differential or unfair treatment in healthcare settings (AOR: 1.56, 95% CI: 1.07– 2.28, p = 0.020). As shown in Table 5, higher odds of moderate-to-severe anxiety symptoms were observed among participants with moderate-to-severe depressive symptoms in comparison to those without (AOR: 26.10, 95% CI: 8.10–84.15, p < 0.001), whereas lower odds were noted among participants attending private tertiary academic institutions than those attending public ones (AOR: 0.30, 95% CI: 0.10–0.88, p = 0.028), and among participants who had never attended boarding school compared with those who had ever (AOR: 0.23, 95% CI: 0.06–0.88, p = 0.032).

**Table 4:** Logistic regression of depressive symptoms and various factors among YSMM (n=224)

| Variable | Category | Unadjusted odds ratios<br>(95% CI) | p value | Adjusted odds ratio<br>(95% CI) | p value |
| --- | --- | --- | --- | --- | --- |
| Gender identity | Cisgender male | Ref |  |  |  |
|  | Transgender female | 2.28 (0.92-5.58) | 0.072 | 2.68 (0.67 – 10.67) | 0.163 |
| Type of academic institution attended | Public | Ref |  |  |  |
|  | Private | 1.58 (0.83 – 2.98) | 0.162 | 0.96 (0.37 – 2.47) | 0.938 |
| Year of study at university/college | 3rd and 4th year | Ref |  |  |  |
|  | 1st and 2nd year | 2.20 (1.08 – 4.47) | 0.029 | 2.79 (1.09 – 7.15) | 0.033* |
| Age at first anal sex with a man | ≥18 years | Ref |  |  |  |
|  | <18 years | 1.52 (0.82 – 2.82) | 0.181 | 1.05 (0.42 – 2.67) | 0.906 |
| Participated in group sex | No | Ref |  |  |  |
|  | Yes | 1.87 (0.88 – 3.95) | 0.102 | 2.71 (0.92 – 7.98) | 0.070 |
| Disclosed same-sex attraction to family members | No | Ref |  |  |  |
|  | Yes | 2.43 (0.74 – 7.98) | 0.143 | 0.26 (0.03 – 2.70) | 0.261 |
| HIV serostatus at time of study | HIV negative | Ref |  |  |  |
|  | HIV positive | 2.95 (1.16 – 7.52) | 0.024 | 4.88 (1.18 – 20.18) | 0.029* |
| Alcohol use based on AUDIT scores | Low-risk (0-7) | Ref |  |  |  |
|  | Hazardous to dependent (≥8) | 3.10 (1.62 – 5.93) | 0.001 | 2.19 (0.85 – 5.63) | 0.105 |
| Ever used non-injectable drugs for non-medical reasons | No | Ref |  |  |  |
|  | Yes | 1.54 (0.82 – 2.87) | 0.172 | 1.09 (0.44 – 2.75) | 0.846 |
| Anxiety symptoms based on GAD-7 scores | None to mild (0-9) | Ref |  |  |  |
|  | Moderate-to-severe (≥10) | 17.47 (7.84 – 38 .94) | <0.001 | 25.63 (8.58 – 76.61) | <0.001* |
| Perceived stress level based on PSS-10 scores | Low (0-13) | Ref |  |  |  |
|  | Moderate-to-high (≥14) | 3.53 (1.43 – 8.71) | 0.006 | 1.71 (0.53 – 5.54) | 0.367 |
| Factor 1: Experienced community/institutional-level discrimination |  | 1.41 (1.07 – 1.85) | 0.014 | 1.24 (0.85 – 1.82) | 0.264 |
| Factor 2: Experienced healthcare-related discrimination |  | 1.26 (0.96 – 1.67) | 0.096 | 1.56 (1.07 – 2.28) | 0.020* |
| Factor 3: Perceived stigma and experienced discrimination in family/religious settings |  | 1.32 (0.99 – 1.75) | 0.055 | 1.20 (0.81 – 1.77) | 0.365 |
| Factor 4: Anticipated stigma in healthcare/social settings |  | 1.15 (0.86 – 1.54) | 0.343 | 0.98 (0.65 – 1.46) | 0.914 |
| Factor 5: Fear and experience of violence |  | 1.58 (1.19 – 2.09) | 0.002 | 1.21 (0.81 – 1.82) | 0.342 |

\*Statistically significant
Note: PCA-derived factor scores were entered as continuous variables. For Factors 1–5, the unadjusted and adjusted odds ratios represent the change in odds associated with a one-unit increase in the respective factor score; no reference category applies. Model diagnostics: Variance Inflation Factors (VIF) analysis indicated no evidence of multicollinearity among exposure variables (all VIFs < 2; tolerance > 0.8; mean VIF = 1.14), supporting the stability of the estimated associations. The Hosmer–Lemeshow goodness-of-fit test indicated adequate model fit $\chi^2 = 7.15$ , p = 0.521).

**Table 5:** Logistic regression of anxiety symptoms and various factors among YSMM (n=226)

| Variable | Category | Unadjusted odds ratios (95% CI) | p value | Adjusted odds ratio (95% CI) | p value |
| --- | --- | --- | --- | --- | --- |
| Age (years) | 21-30 | Ref |  |  |  |
|  | ≥21 | 0.49 (0.25 – 0.96) | 0.038 | 0.43 (0.14 – 1.26) | 0.124 |
| Level of institution attended | College | Ref |  |  |  |
|  | University | 1.92 (0.93 – 3.98) | 0.079 | 2.76 (0.83 – 9.18) | 0.098 |
| Type of academic institution attended | Public | Ref |  |  |  |
|  | Private | 0.32 (0.16 – 0.66) | 0.002 | 0.30 (0.10 – 0.88) | 0.028* |
| Year of study at university/college | 3rd and 4th year | Ref |  |  |  |
|  | 1st and 2nd year | 0.57 (0.26 – 1.23) | 0.154 | 1.37 (0.41 – 4.63) | 0.610 |
| Residence | College/rented hostel | Ref |  |  |  |
|  | With family | 0.48 (0.23 – 1.03) | 0.059 | 0.37 (0.11 – 1.23) | 0.106 |
| Ever attended boarding school | Yes | Ref |  |  |  |
|  | No | 0.52 (0.23 – 1.18) | 0.118 | 0.23 (0.06 – 0.88) | 0.032* |
| Disclosed same-sex attraction to family members | No | Ref |  |  |  |
|  | Yes | 0.43 (0.12 – 1.48) | 0.183 | 0.66 (0.08 – 5.55) | 0.700 |
| Disclosed same-sex attraction to healthcare provider | No | Ref |  |  |  |
|  | Yes | 0.46 (0.20 – 1.04) | 0.065 | 0.47 (0.12 – 1.84) | 0.280 |
| HIV serostatus at time of study | HIV negative | Ref |  |  |  |
|  | HIV positive | 2.29 (0.82 – 6.36) | 0.112 | 0.76 (0.14 – 4.06) | 0.746 |
| Alcohol use based on AUDIT scores | Low-risk (0-7) | Ref |  |  |  |
|  | Hazardous to dependent (≥8) | 1.66 (0.79 – 3.46) | 0.180 | 0.82 (0.27 – 2.50) | 0.728 |
| Depressive symptoms based on PHQ-9 scores | None to mild (0-9) | Ref |  |  |  |
|  | Moderate-to-severe (≥10) | 17.47 (7.84 -38.93) | <0.001 | 26.10 (8.10 – 84.15) | <0.001* |
| Perceived stress level based on PSS-10 scores | Low (0-13) | Ref |  |  |  |
|  | Moderate-to-high (≥14) | 8.10 (1.89 – 34.64) | 0.005 | 3.22 (0.52 – 20.08) | 0.211 |
| Factor 1: Experienced community/institutional-level discrimination |  | 1.37 (1.03 – 1.83) | 0.029 | 1.22 (0.82 – 1.84) | 0.330 |
| Factor 2: Experienced healthcare-related discrimination |  | 0.96 (0.67 – 1.36) | 0.826 | 0.81 (0.48 – 1.38) | 0.445 |
| Factor 3: Perceived stigma and experienced discrimination in family/religious settings |  | 1.24 (0.90 – 1.70) | 0.187 | 1.33 (0.88 – 2.03) | 0.180 |
| Factor 4: Anticipated stigma in healthcare/social settings |  | 1.36 (0.99 – 1.87) | 0.055 | 1.52 (0.97 – 2.39) | 0.070 |
| Factor 5: Fear and experience of violence |  | 1.69 (1.24 – 2.30) | 0.001 | 1.23 (0.79 – 1.90) | 0.357 |

\*Statistically significant
Note: PCA-derived factor scores were entered as continuous variables. For Factors 1–5, the unadjusted and adjusted odds ratios represent the change in odds associated with a one-unit increase in the respective factor score; no reference category applies. Model diagnostics: Variance Inflation Factors (VIF) analysis indicated no evidence of multicollinearity among exposure variables (all VIFs < 2; tolerance > 0.7; mean VIF = 1.17), supporting the stability of the estimated associations. The Hosmer–Lemeshow goodness-of-fit test indicated adequate model fit ( $\chi^2 =$ 205.73, $p = 0.120$ ).

## Discussion

### Key findings

In this study of young sexual minority men (YSMM) enrolled in tertiary academic institutions in Nairobi, Kenya, the estimated prevalence of depressive, anxiety and co-occurring depressive and anxiety symptoms was 23.7%, 16.9% and 12.4%, respectively. Using the same PHQ-9 and GAD-7 cut-offs (≥10), Mbithi *et al.* reported prevalence of 26.0%, 19.1% and 12.5% for depressive, anxiety and co-occurring symptoms among adolescents aged 13-19 years, with prevalence decreasing to 20.6%, 14.5% and 9.1% respectively, for school-going adolescents [52]. These similarities suggest that mental health symptoms constitute an important burden among Kenyan youth generally. However, the distinctive experiences of YSMM including living with HIV and healthcare-related discrimination identified in our study, may represent additional vulnerabilities requiring tailored interventions. Elevated burdens of depression among sexual minority populations have been consistently reported in systematic reviews [2–3]. Similar evidence exists for anxiety and related internalizing conditions among sexual minorities [16]. In sub-Saharan Africa, studies among SMM likewise document considerable depressive symptoms burden within stigmatizing contexts and in settings with constrained access to affirming services [29–31].

After adjustment, predictors for depressive and anxiety symptoms were analytically distinct. Symptoms consistent with moderate-to-severe depression were independently associated with earlier years of study, living with HIV, healthcare-related discrimination, and co-occurring moderate-to-severe anxiety symptoms. Moderate-to-severe anxiety symptoms were independently associated with attending a public tertiary academic institution, boarding school history, and co-occurring moderate-to-severe depressive symptoms. The reciprocal associations between depression and anxiety were strong, indicating substantial comorbidity. Plöderl and Tremblay similarly describe clustering of internalizing disorders among sexual minority populations exposed to chronic stress [16]. Among sexual minority students, stress exposure and perceived safety have also been linked to adverse mental health outcomes [47]. Taken together, these findings underscore the salience of educational stage, institutional conditions, healthcare experiences, and intersecting HIV-related vulnerability for mental health among YSMM in higher education settings.

### Depression-specific interpretation

#### Early academic stage

The independent association between depressive symptoms and being in the first or second year of tertiary education indicates heightened vulnerability during early academic transition. Arnett [17] conceptualizes this period of emerging adulthood as marked by identity consolidation, instability, and reorganization of social relationships, processes that are commonly accompanied by increased emotional distress. Empirical studies among university students consistently report elevated depressive symptoms during the transition from secondary to tertiary education, attributable in part to separation from established support networks and adaptation to unfamiliar academic and social environments [18–19]. For sexual minority youth, these developmental transitions are often compounded by sexual identity development, disclosure processes, and diminished parental support [7,21]. School environments are also recognized as common sites of victimization and exclusion for sexual minority students, with documented implications for mental health [9]. Among YSMM, early engagement in new peer and sexual networks may further heighten exposure to rejection and gender role strain, contributing to psychological distress [10]. Within a vulnerability–stress framework, early academic stage may therefore function as a developmental stressor that interacts with pre-existing psychosocial vulnerabilities to increase the risk of depressive symptoms [6]. Collectively, these findings suggest that the initial years of tertiary education represent a critical period for targeted mental health support among YSMM.

#### Living with HIV

Living with HIV was independently associated with moderate-to-severe depressive symptoms, underscoring the psychological burden of managing chronic illness alongside sexual minority status. Depression among people living with HIV has been widely documented and linked to diagnosis-related distress, treatment demands, concerns about disclosure, and anticipated social rejection [30–31]. In Kenyan SMM populations, depressive symptoms frequently co-occur with HIV-related stigma and broader psychosocial stressors [31]. Among YSMM, HIV-related distress is often intensified by internalized stigma and fears of negative reactions from family members, peers, and healthcare providers [16,29]. Meyer’s Minority Stress Model [4] provides a useful framework for understanding this intersection, conceptualizing co-occurring stigmatized identities as sources of cumulative stress operating across structural, interpersonal, and intrapsychic domains. For YSMM living with HIV in criminalized and socially hostile environments, this convergence of HIV-related and sexuality-related stigma likely amplifies vulnerability to depression. These findings highlight the importance of integrating mental health screening and psychosocial support within HIV services for YSMM.

#### Healthcare-related discrimination

Whereas other stigma, discrimination and violence domains did not retain statistical significance, healthcare-related discrimination was the only domain independently associated with depressive symptoms after adjustment. This PCA-derived component reflected direct experiences of unfair treatment in healthcare settings, including mistreatment by providers, gossip by healthcare workers, and denial of services. The persistence of this association underscores healthcare environments as critical sites of structural vulnerability for YSMM’s mental health. This quantitative finding aligns with qualitative evidence from our earlier work in Nairobi [14,53], where we documented pervasive experiences of stigma, judgment, and service denial among YSMM seeking care in public and academic institution-based health facilities, often resulting in delayed or avoided healthcare engagement. Provider-focused research in the same context further identified moral disapproval, limited training, and attempts to “convert YSMM to heterosexual” as contributors to hostile clinical encounters [54]. Within Meyer’s Minority Stress framework [4], such experiences constitute distal structural stressors that undermine trust in healthcare systems and contribute to psychological distress through repeated institutional invalidation. The attenuation of other stigma domains after adjustment suggests that discrimination within healthcare settings may exert a uniquely potent influence on depressive symptoms in this population. This analytic pattern demonstrates that stigma was examined comprehensively at the item level and rigorously tested in reduced-domain form, strengthening confidence in the adjusted findings. The observed pattern positions healthcare encounters as a particularly salient interface between structural exclusion and individual psychological well-being. It underscores the need for system-level interventions to promote culturally competent, non-judgmental, and affirming care for YSMM.

### Anxiety-specific interpretation

#### Institutional context

Moderate-to-severe anxiety symptoms were independently associated with type of academic institution attended, with lower odds observed among students attending private institutions. Type of institution may reflect differences in student support infrastructure, service accessibility, and perceived safety within educational environments. Cleary *et al*. demonstrated that inadequate institutional support during the transition to higher education is associated with poorer mental health outcomes [19], while Brown *et al*. similarly identified structural and service-level barriers as key impediments to mental health care access among at-risk young people [15]. Among sexual minority students, non-affirming campus climates have been associated with higher levels of stress and anxiety [20–21]. In Nairobi, Mwaniki *et al*. documented that YSMM frequently experience public and academic institution-based facilities as stigmatizing, bureaucratically complex, and difficult to navigate, contributing to psychological distress and avoidance of services [14,53]. Provider-focused research in the same setting further identified moral disapproval and limited competency in sexual minority health needs within institutional environments [54]. Within Meyer’s Minority Stress Model [4], such institutional encounters constitute distal stressors that increase internalizing symptoms through repeated exposure to stigma and diminished perceptions of safety. Taken together, these findings suggest that institutional environments may meaningfully shape anxiety vulnerability among YSMM, with resource availability and affirming practices potentially serving as protective factors.

#### Boarding school history

Anxiety symptoms were also independently associated with boarding school history. This finding should be interpreted cautiously and regarded as hypothesis-generating. Educational environments during adolescence play a critical role in psychosocial development, particularly for sexual minority youth. Toomey and Russell demonstrated that sexual minority students experience significantly higher levels of school-based victimization than their heterosexual peers [9], a pattern reinforced by McGeough and Sterzing’s systematic review of family and institutional victimization [8]. Such adverse exposures have been linked to later depressive and anxiety symptoms among sexual minority young people [3]. Arnett characterizes late adolescence as a sensitive developmental period for identity formation and emotional regulation [17]. Within a vulnerability–stress framework, Goh and Agius describe how early environmental stressors may interact with individual vulnerabilities to increase risk of mental illness [6]. For YSMM, boarding school environments may also reinforce concealment of sexual identity and chronic vigilance – processes Meyer conceptualizes as proximal stress pathways contributing to internalizing distress [4]. Although causal mechanisms cannot be established in this study, prior evidence indicates that adverse school climates among sexual minority youth have lasting implications for mental health [3,9]. Future longitudinal research is needed to clarify how specific boarding school experiences shape anxiety trajectories among YSMM.

### Depression-anxiety comorbidity

A central finding of this study was the extremely strong reciprocal association between depressive and anxiety symptoms, with adjusted odds ratios exceeding 25 in both models. This magnitude indicates substantial co-occurrence of internalizing symptoms among YSMM. Similar clustering of depression and anxiety has been documented among sexual minority populations, where chronic exposure to stigma and stress contributes to overlapping mental health conditions [16]. Reed *et al*. likewise demonstrated that stress exposure and perceived safety are jointly associated with multiple adverse mental health outcomes among sexual minority students [47]. Meyer’s Minority Stress Model [4] provides a useful framework for interpreting this pattern. The model conceptualizes internalizing disorders as interrelated responses to sustained distal and proximal stressors. Within this framework, depression and anxiety are understood as co-occurring manifestations of chronic psychosocial strain rather than isolated conditions. This interpretation aligns with evidence that sexual minority youth exposed to victimization and identity-related stress frequently experience multiple internalizing symptoms simultaneously [3]. Although the cross-sectional design precludes inference regarding directionality, the strength of the observed associations underscores the importance of integrated mental health screening approaches that assess depressive and anxiety symptoms concurrently. Such integrated assessment may be particularly critical for YSMM, who commonly face intersecting stressors related to education, healthcare access, and HIV status [16,31]. From a public health perspective, these findings also resonate with syndemic frameworks, which emphasize how co-occurring psychosocial conditions cluster within structurally marginalized populations to amplify overall disease burden [4,16]. Together, these results highlight the need for comprehensive, integrated mental health services for YSMM in tertiary education settings.

### Implications

The associations between depressive symptoms and earlier year of study, and between anxiety symptoms and type of academic institution (public versus private), underscore the importance of accessible campus-based mental health support for sexual minority youth, particularly during the initial years of tertiary education [55]. Beyond counselling and peer support, interventions should address the broader campus environment, as negative campus climates among sexual minority university students have been associated with greater anxiety and depression symptoms and poorer academic outcomes [56]. Targeted outreach during the transition to tertiary education may help mitigate vulnerability during this critical developmental period, which Arnett characterizes as marked by identity consolidation and emotional instability [17]. Creating affirming educational environments may also serve as an important protective factor for YSMM mental health. Evidence from other settings suggests that cultural-competence training for staff and non-sexual minority students, ally or safe zone programs, explicit anti-discrimination policies, and supportive student structures can improve sexual minority-related knowledge, supportive attitudes, and perceptions of inclusion [57]. However, much of this evidence derives from Western, high-income settings, and its applicability to Kenya requires further evaluation. Future research should therefore examine campus climates across Kenyan public and private institutions, identify contextually appropriate best practices, and assess how institutional policies, staff and student cultural competence, and support structures either promote affirmation or enable discrimination against sexual minority students.

### Limitations

Several limitations warrant consideration. First, the cross-sectional design precludes inference regarding temporal relationships between exposures and outcomes. Second, mental health symptoms and stigma experiences were self-reported, introducing the potential for recall and social desirability bias. Third, the PHQ-9 and GAD-7 are screening instruments rather than diagnostic tools; therefore, although the reported prevalence estimates reflect clinically significant depressive and anxiety symptoms, they should not be interpreted as confirmed clinical diagnoses of depressive or anxiety disorders. Fourth, although multivariate adjustment was applied, residual confounding cannot be excluded, and some estimates were imprecise, as reflected by wide confidence intervals. Finally, while respondent-driven sampling facilitated recruitment of a hidden population, the findings may not be fully generalizable beyond YSMM connected to recruitment networks. Despite these limitations, this study provides important evidence on the burden and predictors of two important mental health conditions among YSMM in Kenya, a population for whom data remain limited.

## Conclusion

Young sexual minority men (YSMM) enrolled in tertiary academic institutions in Nairobi experience a high burden of depressive and anxiety symptoms, with pronounced comorbidity between these conditions. Healthcare-related discrimination emerged as a central structural correlate of depressive symptoms, while early academic stage, HIV status, and institutional context further shaped mental health vulnerability. Together, these findings underscore the need for integrated clinical screening, stigma-reduction within healthcare systems, and targeted campus-based mental health support. Longitudinal research is needed to clarify causal pathways and to inform sustained, structural, and developmentally responsive interventions for YSMM.

## Data Availability

The data analyzed during this study are not publicly available due to the criminalization and stigmatization of some of participants' identities and behaviors. Data may be available from the corresponding author on reasonable request for researchers who meet the criteria for access to confidential data, as determined by the approving ethics committees i.e. University of the Witwatersrand Human Research Ethics Committee Medical (via) and University of Nairobi-Kenyatta National Hospital Ethics and Research Committee (via).

## Acknowledgements

The authors would like to thank Joshua Kimani of Partners for Health and Development in Africa (PHDA) for hosting the data collection at the TRANSFORM study site in Nairobi, Kenya, as well as other staff from PHDA (Rhoda Wanjiru, Monica Okumu, Mary Wanjiru, Hellen Babu, Evelyn Ombunga, Elizabeth Rwenji, Ibrahim Lwingi, Zaina Jama, Pauline Ngurukiri and Laisa Lumumba) for their invaluable support with acquisition of data. We are also grateful to all the study participants without whom the study would not have been possible.

